# Cohort study investigating the natural history and management of sore throat and tonsillitis among adults in UK general practice

**DOI:** 10.64898/2026.02.16.26346374

**Authors:** Samuel Finnikin, James O’Hara, Tom Marshall

## Abstract

**Background:** Recurrent sore throat affects a small minority of adults but can cause substantial morbidity. Evidence to guide tonsillectomy eligibility in adults is limited, and current criteria are extrapolated from paediatric populations. We aimed to describe the epidemiology, management, and prognosis of adult sore throat in UK primary care.

**Methods:** Using CPRD Aurum (2010–2020 adults with a first coded episode of sore throat or tonsillitis were identified and matched to controls. Episode frequency, antibiotic use, ENT referral, and tonsillectomy were analysed. Predictors of recurrent episodes (≥3 in 365 days), referral, and tonsillectomy were assessed using time-to-event, multinomial logistic, and multilevel mixed-effects regression models.

**Findings:** Of 4.45 million adults, 1.70 million (38.3%) had ≥1 episode; most (61.5%) had only one, but 4.1% experienced ≥3 within 1 year. Recurrent episodes were more common in younger females and those from more deprived areas. Only 21,869 patients (0.5% of the exposed cohort) underwent tonsillectomy, and just 25.7% of these met Paradise criteria at any time; conversely, only 13.9% of those meeting criteria underwent surgery. Patients who had a tonsillectomy tended to be younger, female, and from less deprived areas. Pre-tonsillectomy episode rates were unexpectedly low, but the data indicated that individuals with high baseline burden continue to experience elevated episode rates over several years.

**Conclusions:** Recurrent sore throat is uncommon, but those affected face substantial disease burden. Current tonsillectomy patterns are poorly aligned with disease burden and show inequities by deprivation. Earlier identification of adults likely to develop recurrent episodes, and more timely surgical intervention, may improve patient outcomes and the cost-effectiveness of tonsillectomy.

## Background

Sore throat is a common presentation in UK general practice with an incidence of 10 per 1000 population per year. (1) People presenting with sore throat symptoms may be given a specific diagnosis such as tonsillitis or pharyngitis, but the diagnosis is clinical and the aetiology is very seldom confirmed definitively. There is variation in whether people are given specific diagnoses or whether a symptom based ‘sore throat’ diagnosis is made and this has been found to be related to treatment decisions. (2) Tonsillitis, as a cause of sore throat, is unique as there is a surgical treatment available (tonsillectomy) which is indicated if there are recurrent episodes of sore throat caused by tonsillitis. However, the epidemiology of tonsillitis and sore throat in adults is not well described. One Norwegian study quoted the prevalence of self-reported recurrent tonsillitis as 11.7% (3) and the latest consultation rates (2006) results from a large UK primary care dataset were reported as 33.3 and 31.1/1000 patients for tonsillitis (females and males respectively) and 64.0 and 46.5/1000 for sore throat. (4) In most cases (up to 80%) the aetiology of tonsillitis is viral. (5) However, guidelines recommend using clinical criteria and or laboratory tests to target antibiotic treatment to those episodes of tonsillitis most likely to have a bacterial aetiology as antibiotics can confer a small benefit if used accordingly. (6, 7)

Tonsillectomy is undertaken to manage recurrent tonsillitis and it has been shown to improve quality of life in people with recurrent episodes (8) although a 2014 Cochrane review concluded that there was insufficient evidence on the effectiveness on this procedure in adults. (9) The threshold for undertaking tonsillectomy is based on the number of episodes of sore throat and tonsillitis that patients experience over time, and the quality-of-life data suggests that higher thresholds are associated with greater benefits post-tonsillectomy.

Where to set the treatment threshold to maximise the benefit and minimise the harm of tonsillectomy is a matter of debate. UK and international guidance for tonsillectomy eligibility are based on the Paradise criteria and is extrapolated from data in a paediatric population. (10–13) The Paradise criteria specify that patients must have one of; 7 episodes in one year, 5 or more episodes annually in two consecutive years, or 3 or more episodes annually in 3 consecutive years. In a trial published after the Cochrane review, which used the Paradise criteria as the threshold for adult tonsillectomy, the procedure showed a reduction in sore throat days compared with conservative management. (14) However, it is not known whether the Paradise criteria is the most effective way of identifying patients who would benefit most from tonsillectomy. Despite the lack of evidence, these criteria are widely used as commissioning rules governing referral for, and undertaking of, tonsillectomy in the UK.

Patients who have more frequent recurrent sore throats episodes are more likely to benefit from tonsillectomy. However, the natural history of sore throat and tonsillitis in adults is poorly understood. It is unclear whether a history of frequent sore throat episodes in one year (or the Paradise criteria) is a good predictor of sore throat episodes in subsequent years. It is also unclear if other factors such as the patient’s sociodemographic characteristics or other risk factors influence the frequency of occurrence. An understanding of the factors associated with repeated episodes of sore throat would therefore help identify patients who would benefit most from tonsillectomy and provide data to inform their decision-making in relation to tonsillectomy. This research aims to describe the epidemiology, management and prognosis of sore throat and tonsillitis in adults.

## Methods

This retrospective cohort study comprised patients from the Clinical Practice Research Datalink (CPRD) AURUM database. CPRD AURUM contains anonymised primary care records including demographics, coded diagnoses and prescribing data for over 16 million patients from 1,771 practices (around 23.9% of the UK population.) (15)

The data extraction and cohort selection according to study design were facilitated using the data extraction for epidemiological research (Dexter) tool. (16) Patients aged 16 years and over from practices opting into CPRD AURUM were included in the exposed cohort if they had a first, coded episode of tonsillitis or sore throat between 1^st^ January 2010 and 31^st^ December 2020. Clinical codes used are provided in supplementary table S1. All sore throat related diagnoses are included along with symptom-based codes. Tonsillitis codes were separated from all other sore throat diagnoses to allow differentiation in analysis. All non-tonsillitis codes are referred to as ‘sore throat’ in this manuscript.

Patients were eligible for inclusion from the study start date or the earliest of either the practice standardisation date (the date at which the practice data is deemed to be of research quality, based on CPRD algorithm) or date the patient registered with the practice plus 3 years, until the earliest of: study end date, end of practice data, or death. The date of the first incident diagnosis of tonsillitis or sore throat following the three-year run-in period was defined as the index date. The 3-year run-in period was chosen so that the index date was not necessarily the patients first episode of tonsillitis/sore throat. This allowed for a baseline episode rate to be established in the first year of follow up, and both retrospective and prospective analysis from the index date (supplementary figure S1). Patients were excluded if they left the database within 1 year and 30 days of this as a minimum of 1 year follow up was required and the follow up period started 30 days after the index date (to allow for the episode to resolve). Patients were also excluded if they had previous tonsillectomy or an indication for tonsillectomy other than recurrent sore throat (i.e. tonsillar malignancy) at any time before or during the study period.

A matched comparison group was created consisting of age, sex and practice matched patients who did not have a coded entry for tonsillitis or sore throat in the study period. Patients were matched at a 1:2 ratio (exposed to unexposed).

All coded episodes of tonsillitis or sore throat were identified (herein referred collectively as ‘episodes’). Episodes were defined as any code for sore throat including pharyngitis, tonsillitis or sore throat symptoms at the time of consultation or when there was a healthcare visit recording of sore throat or tonsillitis (including out-of-hours GP, walk-in primary care centre or emergency department visit). Ascertainment of episodes was performed recursively with every episode including a 30-day grace period before a further episode could be identified to allow for full remission of that episode. This time period was chosen as a clinically realistic time to allow symptoms to completely resolve. Covariates included were sex, age, ethnicity, body mass index (BMI) smoking status, geographic region and deprivation (index of multiple deprivation (IMD) quintile). These were chosen as the variables most likely to be associated with sore throat episodes and subsequent treatment decisions. A ‘missing’ category was used for missing covariate data as these were assumed not to be missing at random.

All prescriptions for antibiotics used in the treatment of sore throat and tonsillitis (phenoxymethylpenicillin, amoxicillin, clindamycin, clarithromycin and erythromycin) were obtained. Efforts were made to distinguish between repeat prescriptions (as these would be for other indications (e.g. prophylactic antibiotics)) and acute prescriptions (used in the treatment of acute infections). The coded prescription duration was deemed to be unreliable so identification of repeat prescriptions was done by: [1] calculating expected duration by using the total dose and usual daily dose and removing those with a calculated duration of >14 days [2] removing prescriptions for patients who had >40 prescriptions of the same antibiotic, and [3] identifying regular prescriptions by looking at the variance of the prescription interval for people prescribed >10 of the same antibiotic and removing those that appeared to be a regular (e.g. monthly) prescription. Repeat prescriptions were removed from analysis.

Referrals to Ear, Nose and Throat (ENT) specialists were identified as outcome variables along with tonsillectomy codes. Referrals were subclassified into those within 180 days of a sore throat episode as these would be more likely to be causally linked to the sore throat episode.

### Statistical analysis

The cohort was described in terms of sociodemographic variables. The pattern of sore throat episodes was described using summary statistics and the temporal relationship with the index date. Concurrent antibiotic prescribing was described along with ENT referrals and tonsillectomies performed. The population who underwent tonsillectomy was described and their episode rate pre- and post-tonsillectomy was calculated.

Patients who met the Paradise criteria for tonsillectomy at any time (prior to or after the index date) were identified. The proportion of these who underwent tonsillectomy were described, along with the time to tonsillectomy and episodes post-tonsillectomy. Exploratory analysis was undertaken using hypothetical tonsillectomy criteria to examine the potential benefit of different thresholds for tonsillectomy. These were set at 3, 4, 5, 6, or ≥7 episodes in any 365-day period, and episodes after reaching this threshold were described along with potential benefits (in terms of sore throat episodes avoided excluding post-surgical pain) that could be seen if tonsillectomy were to have been performed. To explore the pattern of recurrent episodes, patients with at least one episode in the baseline year (i.e. the exposed cohort) who had at least 4 years of follow up and who did not undergo tonsillectomy were categorized according to the number of episodes they experienced in the baseline year. The episodes experienced in the preceding and following three years were described according to these categories.

The factors associated with recurrent episodes were described using a time to event analysis considering multiple events from the index episode using the Anderson-Gill Cox model. (17) Predictor variables were the demographic variables above, along with the number of episodes in the three years before the index episode. Three years was chosen as the current tonsillectomy criteria considers this timeframe. Multinomial logistic regression analyses were undertaken looking at the predictors of ENT referral (ever, and within 180 days of an episode), tonsillectomy and the chances of patients having three episodes in one year. Multilevel mixed-effect linear regression explored the predictors of the episode rate.

Three episodes in one year was chosen as a predictor variable as this is the minimum rate of recurrent episodes that could lead to tonsillectomy under current commissioning guidance. GP practice ID was applied as a random effect in all models.

To establish whether the results would be affected by potentially uncoded episodes, sensitivity analysis was undertaken to identify episodes that were not coded but instead identified through an acute prescription of phenoxymethylpenicillin (penV) since this antibiotic is almost exclusively used for this indication when prescribed acutely.

The study protocol (ID 24_004512) was approved by the CPRD expert review committee.

## Results

A total of 4,453,538 patients were identified from 1771 practices (see STROBE diagram supplementary figure S2.) The exposed cohort (patients who had at least one episode) comprised 1,704,704 patients (38.3%) and, compared to unexposed, these patients were statistically more likely to be female (62.7% vs 58.7%, p value <0.0001) younger (mean age 40.3 vs 41.9 years, p<0.001) and of white ethnicity (71.9% vs 67.5%, p<0.001) (table 1.) The mean observation period (including the run-in period) was 8.5 years. Patients in the exposed cohort had significantly more consultations per year than the unexposed cohort (19.7 vs 13.3 P<0.001). Overall, 3,076,128 episodes were recorded: two thirds were sore throat diagnoses (1,988,376 (64.6%). 329,517 (10.7%) of episodes occurred pre-index, and 1,041,907 (33.9%) post-index. The Kernel density plot shows the distribution of episodes around the index date (supplementary figure S3).

**Table 1:**
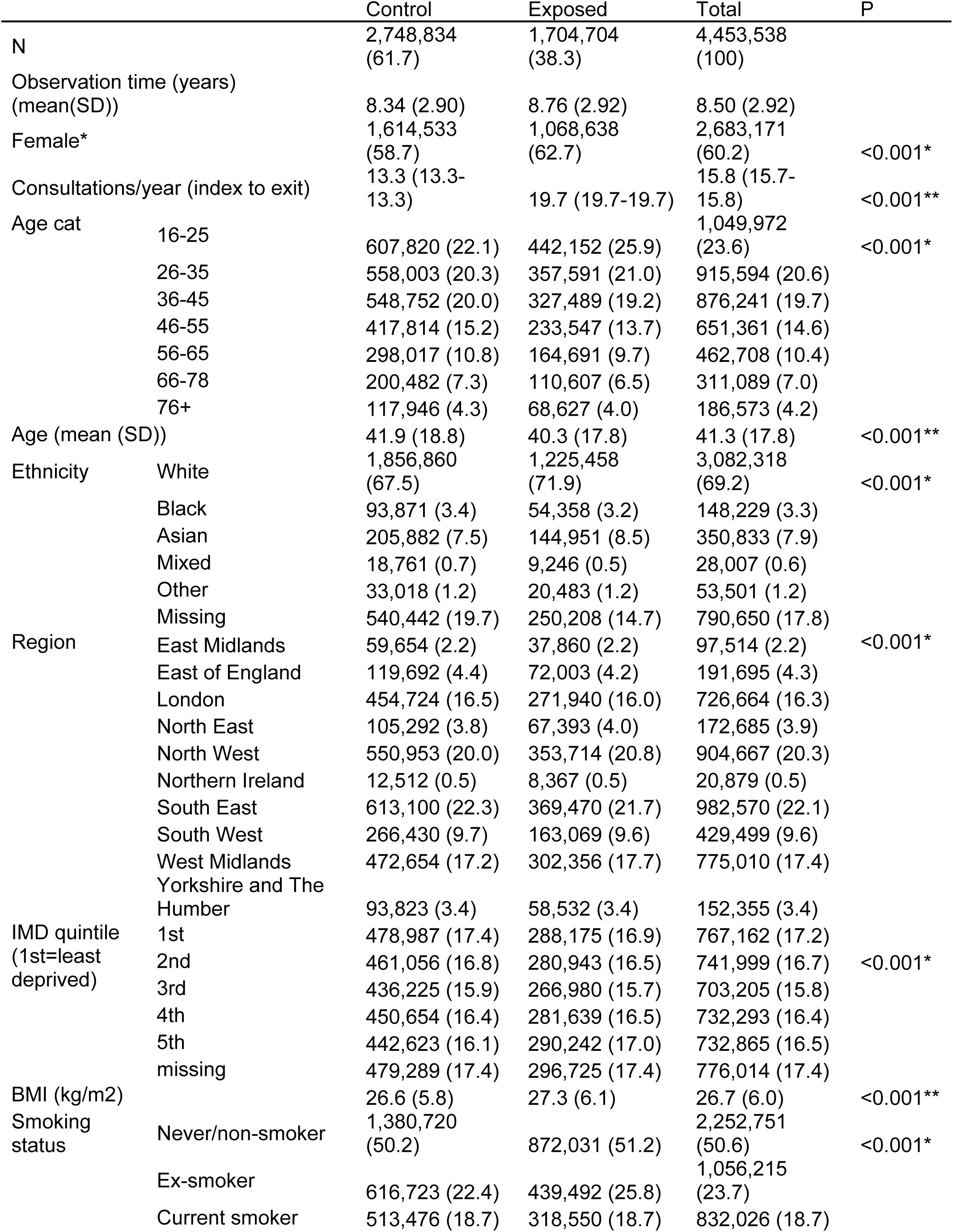
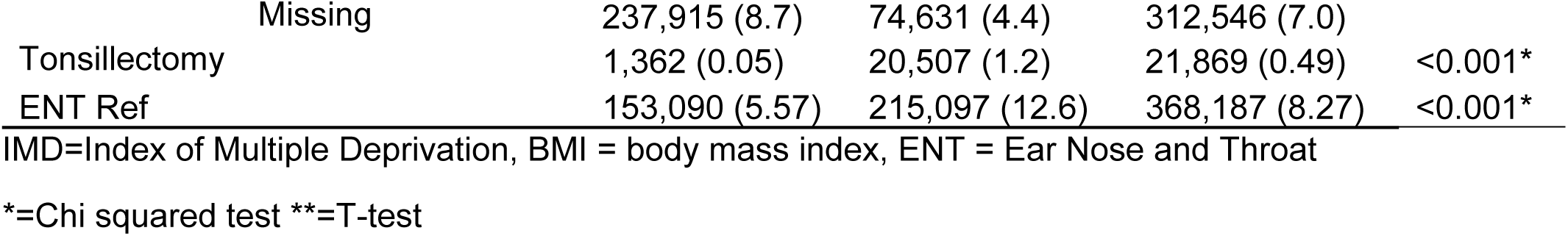
Cohort Characteristics.

|  |  | Control | Exposed | Total | P |
| --- | --- | --- | --- | --- | --- |
| N |  | 2,748,834<br>(61.7) | 1,704,704<br>(38.3) | 4,453,538<br>(100) |  |
| Observation time (years)<br>(mean(SD)) |  | 8.34 (2.90) | 8.76 (2.92) | 8.50 (2.92) |  |
| Female* |  | 1,614,533<br>(58.7) | 1,068,638<br>(62.7) | 2,683,171<br>(60.2) | <0.001* |
| Consultations/year (index to exit) |  | 13.3 (13.3-13.3) | 19.7 (19.7-19.7) | 15.8 (15.7-15.8) | <0.001** |
| Age cat | 16-25 | 607,820 (22.1) | 442,152 (25.9) | 1,049,972<br>(23.6) | <0.001* |
|  | 26-35 | 558,003 (20.3) | 357,591 (21.0) | 915,594 (20.6) |  |
|  | 36-45 | 548,752 (20.0) | 327,489 (19.2) | 876,241 (19.7) |  |
|  | 46-55 | 417,814 (15.2) | 233,547 (13.7) | 651,361 (14.6) |  |
|  | 56-65 | 298,017 (10.8) | 164,691 (9.7) | 462,708 (10.4) |  |
|  | 66-78 | 200,482 (7.3) | 110,607 (6.5) | 311,089 (7.0) |  |
|  | 76+ | 117,946 (4.3) | 68,627 (4.0) | 186,573 (4.2) |  |
| Age (mean (SD)) |  | 41.9 (18.8) | 40.3 (17.8) | 41.3 (17.8) | <0.001** |
| Ethnicity | White | 1,856,860<br>(67.5) | 1,225,458<br>(71.9) | 3,082,318<br>(69.2) | <0.001* |
|  | Black | 93,871 (3.4) | 54,358 (3.2) | 148,229 (3.3) |  |
|  | Asian | 205,882 (7.5) | 144,951 (8.5) | 350,833 (7.9) |  |
|  | Mixed | 18,761 (0.7) | 9,246 (0.5) | 28,007 (0.6) |  |
|  | Other | 33,018 (1.2) | 20,483 (1.2) | 53,501 (1.2) |  |
|  | Missing | 540,442 (19.7) | 250,208 (14.7) | 790,650 (17.8) |  |
| Region | East Midlands | 59,654 (2.2) | 37,860 (2.2) | 97,514 (2.2) | <0.001* |
|  | East of England | 119,692 (4.4) | 72,003 (4.2) | 191,695 (4.3) |  |
|  | London | 454,724 (16.5) | 271,940 (16.0) | 726,664 (16.3) |  |
|  | North East | 105,292 (3.8) | 67,393 (4.0) | 172,685 (3.9) |  |
|  | North West | 550,953 (20.0) | 353,714 (20.8) | 904,667 (20.3) |  |
|  | Northern Ireland | 12,512 (0.5) | 8,367 (0.5) | 20,879 (0.5) |  |
|  | South East | 613,100 (22.3) | 369,470 (21.7) | 982,570 (22.1) |  |
|  | South West | 266,430 (9.7) | 163,069 (9.6) | 429,499 (9.6) |  |
|  | West Midlands | 472,654 (17.2) | 302,356 (17.7) | 775,010 (17.4) |  |
|  | Yorkshire and The<br>Humber | 93,823 (3.4) | 58,532 (3.4) | 152,355 (3.4) |  |
| IMD quintile<br>(1st=least<br>deprived) | 1st | 478,987 (17.4) | 288,175 (16.9) | 767,162 (17.2) |  |
|  | 2nd | 461,056 (16.8) | 280,943 (16.5) | 741,999 (16.7) | <0.001* |
|  | 3rd | 436,225 (15.9) | 266,980 (15.7) | 703,205 (15.8) |  |
|  | 4th | 450,654 (16.4) | 281,639 (16.5) | 732,293 (16.4) |  |
|  | 5th | 442,623 (16.1) | 290,242 (17.0) | 732,865 (16.5) |  |
|  | missing | 479,289 (17.4) | 296,725 (17.4) | 776,014 (17.4) |  |
| BMI (kg/m2) |  | 26.6 (5.8) | 27.3 (6.1) | 26.7 (6.0) | <0.001** |
| Smoking<br>status | Never/non-smoker | 1,380,720<br>(50.2) | 872,031 (51.2) | 2,252,751<br>(50.6) | <0.001* |
|  | Ex-smoker | 616,723 (22.4) | 439,492 (25.8) | 1,056,215<br>(23.7) |  |
|  | Current smoker | 513,476 (18.7) | 318,550 (18.7) | 832,026 (18.7) |  |
|  | Missing | 237,915 (8.7) | 74,631 (4.4) | 312,546 (7.0) |  |
| Tonsillectomy |  | 1,362 (0.05) | 20,507 (1.2) | 21,869 (0.49) | <0.001* |
| ENT Ref |  | 153,090 (5.57) | 215,097 (12.6) | 368,187 (8.27) | <0.001* |

IMD=Index of Multiple Deprivation, BMI = body mass index, ENT = Ear Nose and Throat
\*=Chi squared test \*\*=T-test

The median number of episodes experienced by people in the exposed group was 1 (IQR 1-2) with 1,047,938 (61.5%) experiencing just one episode (the index episode). The maximum number of episodes was 58 in one individual. supplementary table S2 shows how many individuals experienced each number of episodes. Regression modelling for episode rate shows a higher coefficient for females over males (0.015, 95%CI 0.015-0.016), and Asian ethnicity over all others as well as a decreasing trend with age and an increasing trend with increasing deprivation (table 2). Out of the exposed cohort, 70,214 (4.1%) experienced at least 3 episodes in any 365d period; the odds of which were highest in the youngest age group, females and those with Asian ethnicity and there were higher odds in areas of higher deprivation, with the most deprived quintile having an OR of 1.16 (95%CI 1.10-1.22) compared with the least (table 2).

**Table 2:**
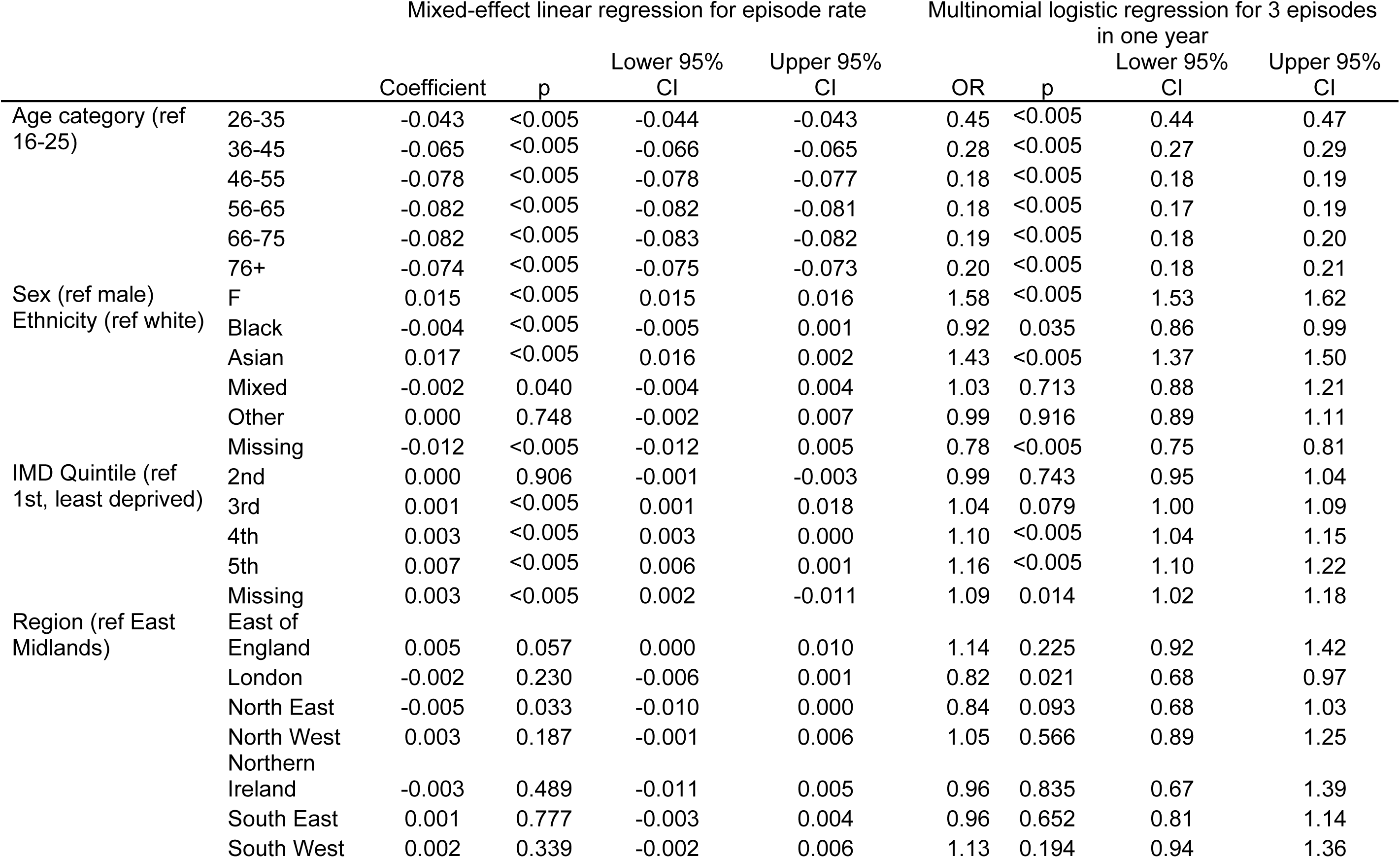
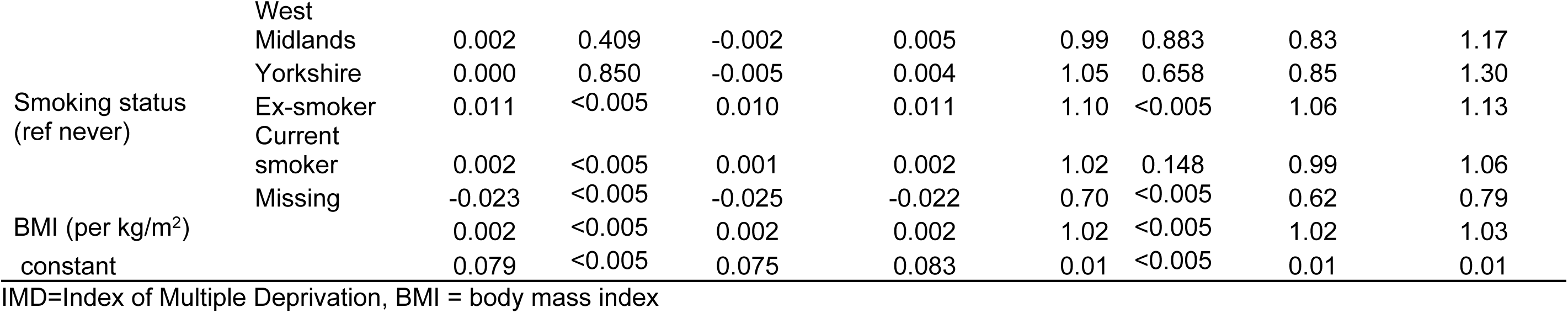
Regression modelling for episode rate and the odds of a patient experiencing 3 or more episodes in any one year.

Taking the 1,257,480 people in the exposed group who had at least 4 years of follow up who did not have a tonsillectomy, the pattern of recurrent episodes is shown in supplementary table S3. A significant proportion of people with a high number of episodes in the baseline year go on to have recurrent episodes in the subsequent years and have a high burden of disease, but equally there is evidence of regression to the mean post baseline year with the proportion of people having no episodes 2 years after baseline being between 84% (for 1 episode in baseline year) and 31% (>6 episodes in baseline year). Figure 1 shows the mean number of episodes per year categorised by the number of episodes in the baseline year.

**Figure 1:**
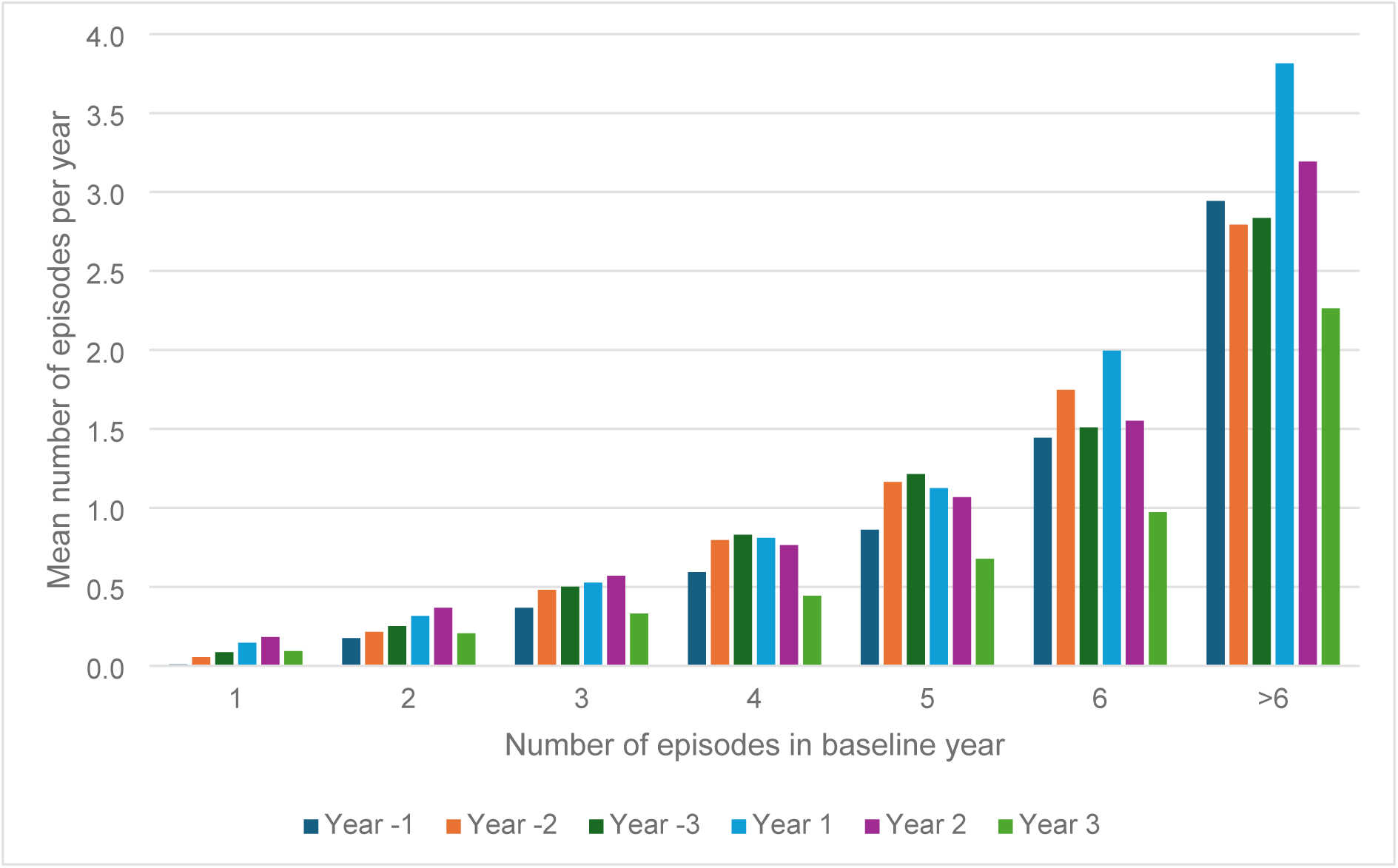
Mean number of episodes per year categorised by number of episodes in baseline year

The number of people reaching a theoretical tonsillectomy threshold of 3, 4, 5, 6, or >6 episodes in any 365-day period are shown in table 3 along with the number of episodes that these patients experienced after reaching these thresholds. This allows for a theoretical reduction in episodes that could be achieved by tonsillectomy (if it was performed immediately after the threshold was reached and not including post-surgical pain) assuming an annual post-tonsillectomy episode rate of 0.1 per person per year. At the current, and highest, 365-day threshold of 7 or more episodes, a tonsillectomy is predicted to prevent 3.9 episodes per person over a 7-year period.

**Table 3:**
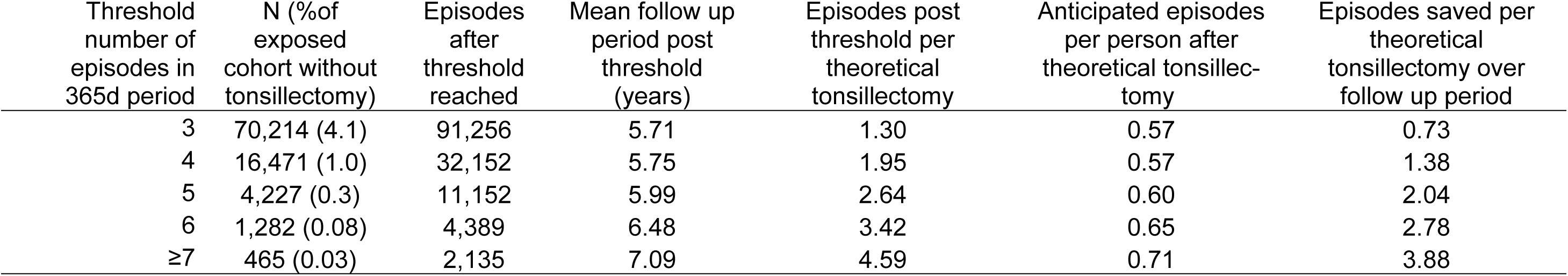
Number of people reaching theoretical 365-day episode thresholds and the potential episodes saved through tonsillectomy for different thresholds.

| Threshold number of episodes in 365d period | N (%of exposed cohort without tonsillectomy) | Episodes after threshold reached | Mean follow up period post threshold (years) | Episodes post threshold per theoretical tonsillectomy | Anticipated episodes per person after theoretical tonsillectomy | Episodes saved per theoretical tonsillectomy over follow up period |
| --- | --- | --- | --- | --- | --- | --- |
| 3 | 70,214 (4.1) | 91,256 | 5.71 | 1.30 | 0.57 | 0.73 |
| 4 | 16,471 (1.0) | 32,152 | 5.75 | 1.95 | 0.57 | 1.38 |
| 5 | 4,227 (0.3) | 11,152 | 5.99 | 2.64 | 0.60 | 2.04 |
| 6 | 1,282 (0.08) | 4,389 | 6.48 | 3.42 | 0.65 | 2.78 |
| ≥7 | 465 (0.03) | 2,135 | 7.09 | 4.59 | 0.71 | 3.88 |

Antibiotics were prescribed for 81.9% of tonsillitis episodes and 39.7% of sore throat episodes. Phenoxymethylpenicillin was the most commonly prescribed antibiotic comprising 81.5% of prescriptions for tonsillitis and 28.2% of prescriptions for sore throat (supplementary table S4.) Overall, 368,187 (8.3%) of patients were referred to ENT, 12.6% of the exposed group and 5.6% of the control group. Of these referrals, 81,616 were within 180 days of a sore throat episode representing 4.8% of the exposed group. 21,869 (0.5%) of the whole cohort underwent tonsillectomy, most of whom (93.8%) were in the exposed group (supplementary table S5). 242 people met the Paradise criteria at baseline (Paradise positive at index) and 40,427 people (2.4% of exposed population) were Paradise positive at some stage during the study period. Of the 21,869 people who underwent tonsillectomy, 5,629 (25.7%) were Paradise positive at any time. Of those patients who met the Paradise criteria at any time, 14,481 (35.8%) were referred to ENT and 5,629 (13.9%) had a tonsillectomy (figure 2).

**Figure 2:**
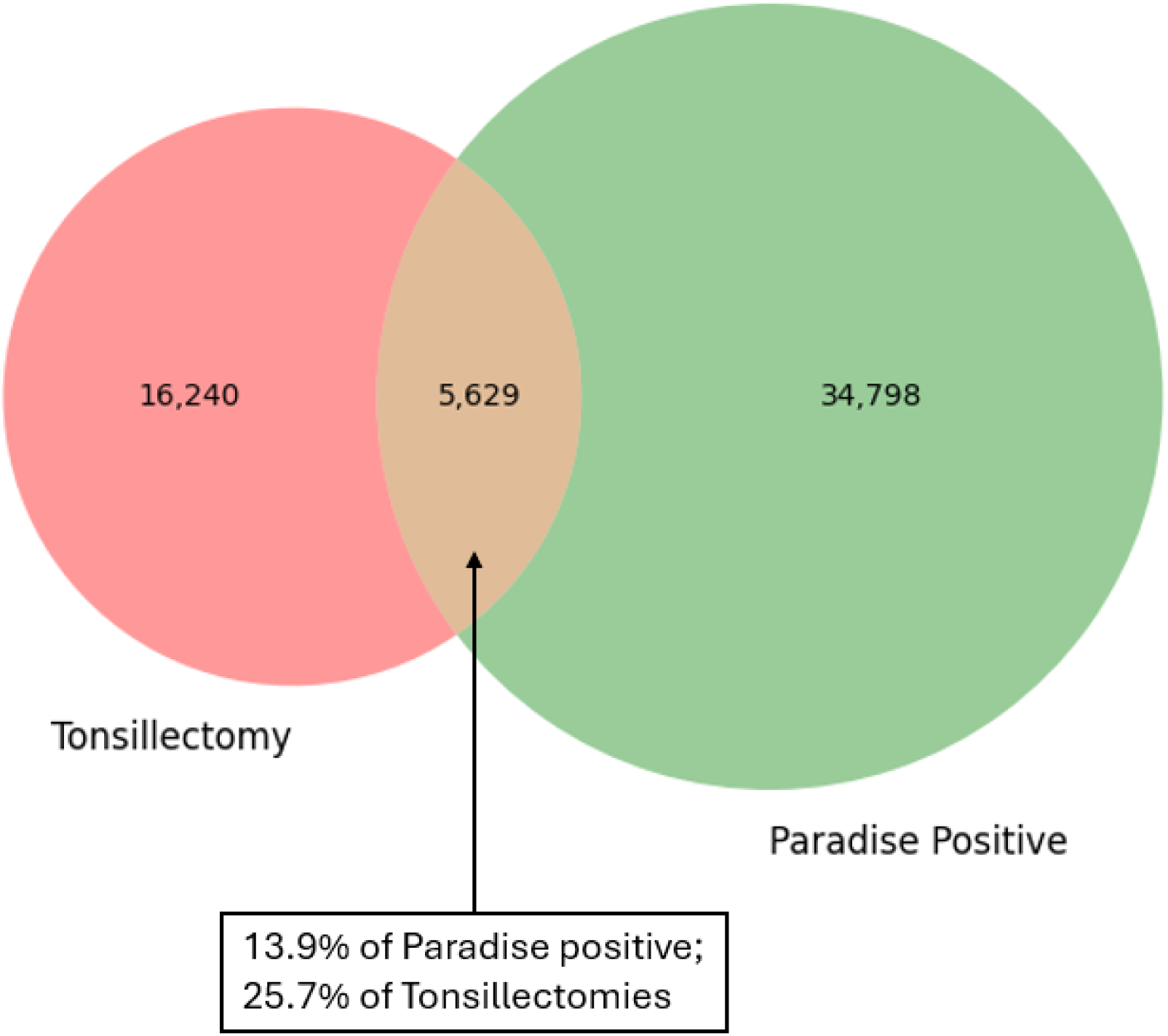
Venn diagram showing relationship between patients who reached the Paradise criteria at any time during the cohort, and patients who underwent tonsillectomy.

The median age of the 21,869 people who had a tonsillectomy was 23.2 years (IQR 18.0-33.8) and 71.0% were female. Pre-tonsillectomy, this group experienced a mean of 3.5 episodes at a rate of 0.7 episodes per person year which reduced to 0.1 episodes per person year post-tonsillectomy. Linear regression modelling for the predictors of tonsillectomy is shown in supplementary table S6. Being Paradise positive was the most significant predictor (OR 27.5, 95% CI 26.3-28.8) but there was also a sharp reduction in OR in all age groups compared with the baseline group (16-25 years) and females were more likely to undergo tonsillectomy (OR 1.18. 95%CI 1.13-1.23). There was no significant regional variation across England, but patients in Northern Ireland were more likely to have a tonsillectomy (OR 1.8, 95%CI 1.3-2.5.) Notably, the odds of undergoing tonsillectomy were reduced with increasing levels of deprivation down to an OR of 0.80 (0.74-0.85) at the highest level of deprivation compared with the lowest. A corresponding deprivation gradient in the odds of referral to ENT was also observed, although not as marked and this gradient was not present when looking only at referrals within 180 days of referral (supplementary table S7).

The results of the survival analysis are shown in supplementary table S8. The number of episodes in the preceding 3 years is the greatest predictor of a future episode along with younger age and being female (figure 3).

**Figure 3:**
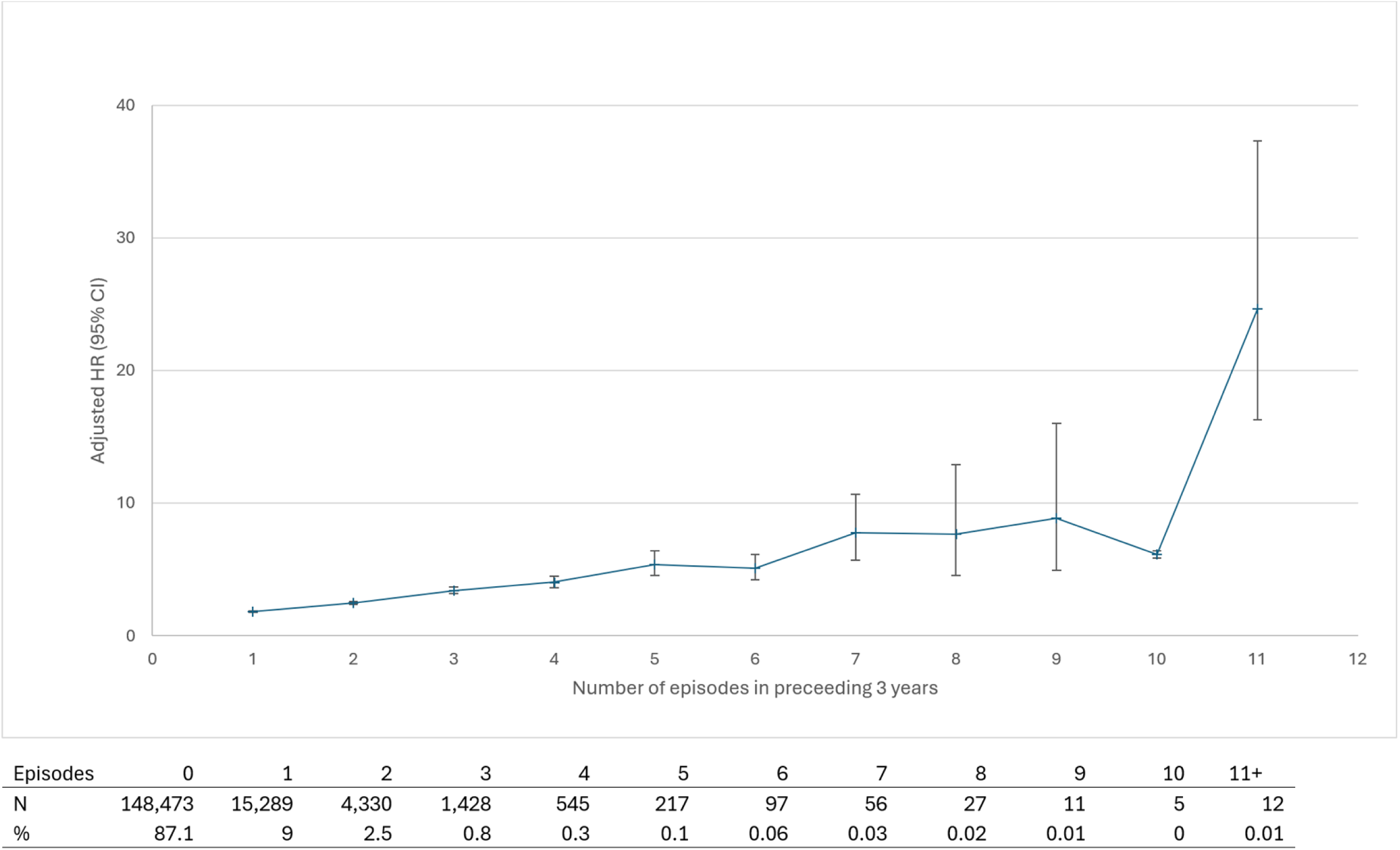
Effect of number of episodes in the preceding 3 years on the hazard of a further episode occurring

Sensitivity analysis including episodes defined by acute prescriptions of penV increased the number of episodes identified by 489,322 (supplementary table S9). In these newly identified episodes, many were in individuals with a high number or recurrent episodes which, despite efforts to try and exclude them, suggested these could represent repeat prescriptions rather than acute prescriptions and therefore not be indicative of a sore throat episode. Given the relatively low prevalence of recurrent episodes, these extra, and potentially mis-ascertained, cases had a significant effect on the modelling. Therefore, it was decided that the additional cases ascertained by antibiotic prescribing reduced the reliability of the results and did not add sufficient data to be useful. However, the antibiotic defined episodes did allow an exploratory analysis (supplementary material S1) suggesting that only 6.8% of episodes presenting to primary care were uncoded. Therefore, using the coded episodes alone (thereby not including antibiotic defined and uncoded episodes) is likely to represent, at a conservative estimate (assuming that all antibiotic defined episodes are accurate,) 76% of all episodes that present to primary care.

## Discussion

Recurrent sore throat symptoms are rare, but people who have multiple episodes often go on to have significant disease burden over the course of a few years. However, regression to the mean seems to occur so that over longer time periods the chances of multiple episodes over a year are low. Younger, female patients have highest chance of recurrent episodes and there is an increasing trend with increasing deprivation. The odds of tonsillectomy decrease with increasing deprivation highlighting a disparity between disease burden and surgical management which does not appear to be fully explained by referral behaviour.

Antibiotics are frequently used in the management of tonsillitis, but less so in other sore throat diagnoses. Tonsillectomy was uncommon, and most people who underwent tonsillectomy did not meet the Paradise criteria according to the primary care record. Most people who would be eligible for tonsillectomy did not get referred to ENT or subsequently undergo the procedure. It is notable that the rate of episodes in the people who underwent tonsillectomy was low with an average of less than one episode per year. Qualitative research amongst UK GPs showed an awareness, and consideration, of referral thresholds and only generally refer if patients make a specific request and they meet the referral threshold. (18) The low episode rate prior to tonsillectomy may be due to underreporting of episodes in the primary care record either due to lack of coding (which we estimate to be up to a quarter of episodes), self-management of episodes, or episodes managed outside of general practice (for example in walk-in-centres) that were not coded. It could also be that the crude episode count doesn’t capture the burden of disease patients are experiencing, and patients may request referral when their disease burden is high despite their number of episodes being relatively low. It is also possible that patients suffered a significant number of episodes >3 years pre-index date which would not have been captured in our methods.

### Comparisons with existing literature

A retrospective chart analysis of 44 Israeli adults attending a military otolaryngology unit who underwent tonsillectomy showed a mean of 1.89 episodes of tonsillitis in the year before surgery, with only one patient meeting the Paradise criteria. (19) This small cohort shows a similar disconnect between the Paradise criteria and tonsillectomy and a lower-than-expected pre-tonsillectomy episode rate, albeit higher than we observed. The 1,122 adults who underwent surgery in a Swedish cohort had a very similar demographic profile to that reported in this study (68.6% female, mean age at tonsillectomy 25.1 years). (20) In this cohort, patients had a mean of 1.4 episodes per year in the 2 years prior to tonsillectomy, again higher than the 0.7 episodes per year that we observed but this may be due to longer period of observation in our study (5 years pre-tonsillectomy.) This cohort observed a reduction in medical care visits post-tonsillectomy of -1.3/year, but a matched control group saw a reduction in visits of -1.2/year resulting in a benefit in the tonsillectomy group of -0.1 visits per year. The authors suggest that regression to the mean as an explanation for this observation, a finding that is supported by a survey of patients on a tonsillectomy waiting list where a significant reduction in symptoms was overserved whilst waiting for tonsillectomy over a 2-year period. (21) These observations are consistent with the pattern of recurrent episodes we have reported.

Another Swedish cohort (22) found the proportion of tonsillitis episodes treated with antibiotics was 73% which is lower than the proportion in our cohort (82%) but sore throat episodes were not reported in the Swedish cohort so there may be some differences in classification which could explain this given that sore throats are less likely to be treated with antibiotics (39.7%). Prescribing rates for sore throat and tonsillitis in the UK have been reported to be higher than in our cohort, but these data are older and prescribing rates have decreased over time. (4, 23, 24)

When the indication for tonsillectomy was examined in a similar cohort of children in the UK, only 11.7% had evidence-based indications (mainly meeting the Paradise criteria) which is lower than we found in the adult population. (25) It was also reported that 5.1% of children who underwent tonsillectomy had no recorded episodes of sore throat/tonsillitis or other indications, a similar proportion to the 6% of our cohort who had no episodes reported prior to tonsillectomy.

### Strengths and limitations

This primary care based cohort study comprised a large adult population over a long study period allowing for the natural history of sore throat in adults to be described in more detail than has been previously reported. The diagnostic coding of sore throat and tonsillitis episodes, whilst not complete, appears to capture a good proportion of episode that present to medical care, a finding supported by the examination of the prescribing data (which is comprehensive).

Not all episodes of tonsillitis and sore throat will have been recorded in the coded data and therefore our analysis does not capture the full burden of disease. Some people will have presented to general practice and been managed without a code being generated; we have estimated that this is a small proportion of the total number of episodes. Others may have presented to walk-in-centres, out-of-hours GPs or emergency departments and these episodes, and any associated prescribing, may not have been subsequently coded in the GP record. However, with general practice seeing over 10 times more patients per year than emergency departments, (26) the contribution of episodes managed in emergency departments is likely to be small. Similarly, out of hours general practice manages much fewer patients than in hours general practice (5.8m in 2013/14 compared with around 300 million in hours) (27) meaning the contribution to total episodes is likely to be small. Finally, a proportion of patients will self-manage their sore throat symptoms (28) and these episodes will be unrecorded. However, when considering indication for tonsillectomy, as these episodes are likely to represent episodes that are less severe and have lower impact on a patient’s functioning, these would not count towards meeting the Paradise criteria. If self-reporting of self-managed sore throat episodes are being used in decision making for tonsillectomy, it is important to note that the accuracy of this information is likely to be subject to considerable bias. (29, 30) It is also worth noting that deprivation may not affect the propensity of patients to present to general practice with sore throat symptoms (28) so consulting behaviour alone is unlikely to explain the observed associations with deprivation.

### Implications for practice and research

This is the first large scale cohort of adult patients with sore throat which therefore expands our understanding of the natural history of this condition. Our findings indicate that the burden of recurrent sore throat falls on a small proportion of the population who experience tonsillitis and sore throat, but there appears to be a period of relatively intense and severe disease in some people. It is possible that identifying these patients earlier in the disease process, and intervening quickly with a prompt tonsillectomy, may be the optimal strategy to reduce the morbidity from tonsillitis and maximise the benefit from surgery. Similarly, if people have been experiencing recurrent episodes over a few years, then the benefit they could realise from surgery may be reduced as the chances of experiencing a high number of episodes in a year tends to reduce after a few years (see supplementary table S3). These insights could inform potential strategies for future research into optimal tonsillectomy threshold and timing.

In practice, there is a mismatch between those who are currently thought to be most likely to benefit from tonsillectomy (those who meet the Paradise criteria) and those who are undergoing surgery. This is perhaps surprising given the widespread awareness of these criteria through commissioning guidelines a finding that warrants further enquiry. There is, perhaps, potential to use the electronic patient record, along with accurate coding, to automatically identify, and share across primary and secondary care, those patients who meet the Paradise criteria to improve patient selection for tonsillectomy.

The discrepancy between the disease burden and tonsillectomy rates by deprivation gradient is noteworthy. Whether this is due to differences in health seeking behaviour, referral practices, surgical decision making, commissioning or access to primary or secondary care is not clear. Further research could identify the factors contributing to this inequity and inform solutions.

## Conclusion

This research gives insight into the epidemiology of recurred sore throat in the UK adult population that may provide potential strategies for improved targeting of surgical intervention. A small proportion of the population experience recurrent sore throat, but intervening early in this population could improve the clinical and cost-effectiveness of tonsillectomy. Currently the population undergoing tonsillectomy does not appear to be well aligned with the population experiencing the most burden of disease, and there is an unjustified deprivation inequality. Our findings could be used to investigate new strategies to identify adults more likely to experience recurrent sore throats and the potential of more timely intervention to reduce disease burden and increase the cost-effectiveness of tonsillectomy.

## Data Availability

Primary data is not available through the authors due to data sharing restrictions but is available to researchers through CPRD)https://www.cprd.com/) subject to approvals. Code lists used in data extraction and processing are available on request from the corresponding author.

## Declarations

### Ethics approval and consent to participate

The study protocol was reviewed and approved by CPRD (Protocol 24_004512) who hold ethical approval (21/EM/026) for use of data without individual patient consent

### Consent for publication

No specific consent was sought or required

### Availability of data and materials

Primary data is not available through the authors due to data sharing restrictions but is available to researchers through CPRD (subject to approvals). Code lists used in data extraction and processing are available on request from the corresponding author.

### Authors’ contributions

◦ Conceptualization -SF, TM, JOH
◦ Data curation - SF
◦ Formal analysis - SF
◦ Investigation - SF
◦ Methodology – SF, TM, JOH
◦ Writing – original draft - SF
◦ Writing – review and editing SF, TM, JOH

## Acknowledgements

None

## Supplementary material

**Supplementary figure S1:**
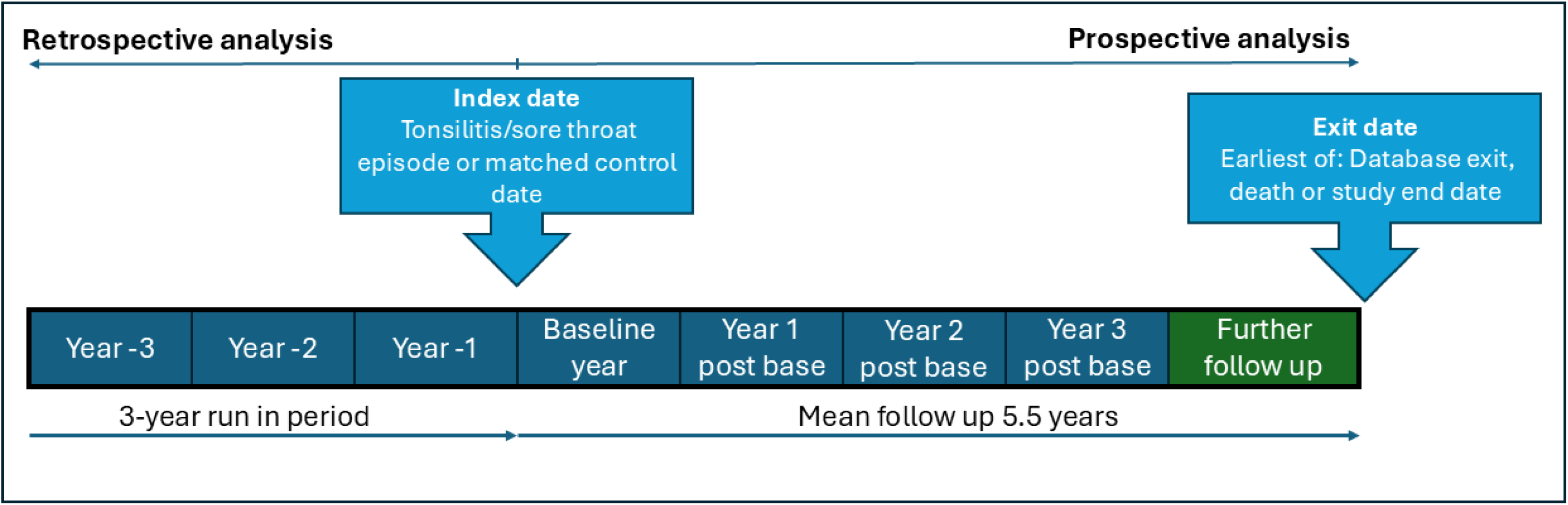
Schematic of the study

**Supplementary figure S2:**
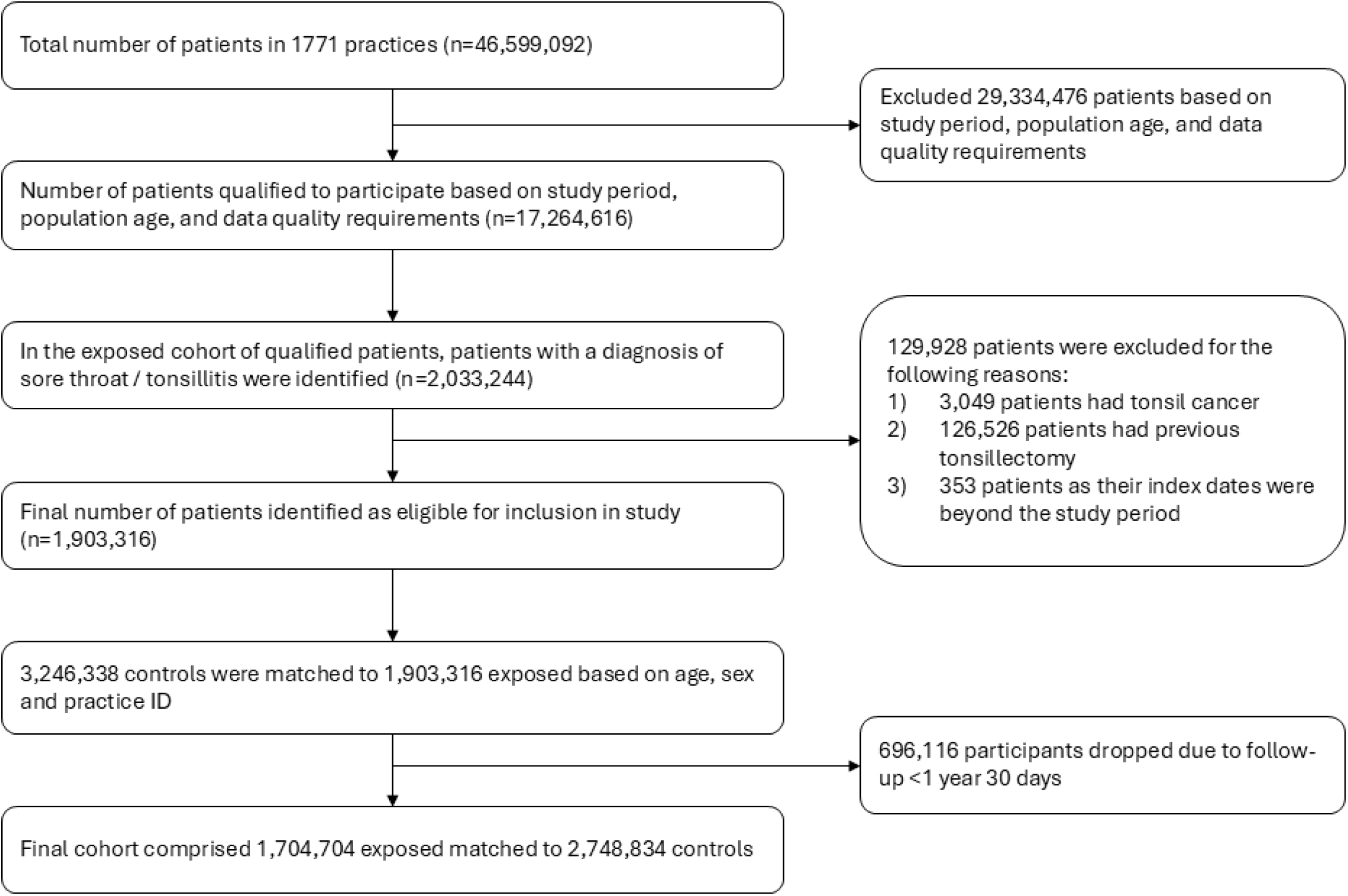
STROBE diagram

**Supplementary figure S3:**
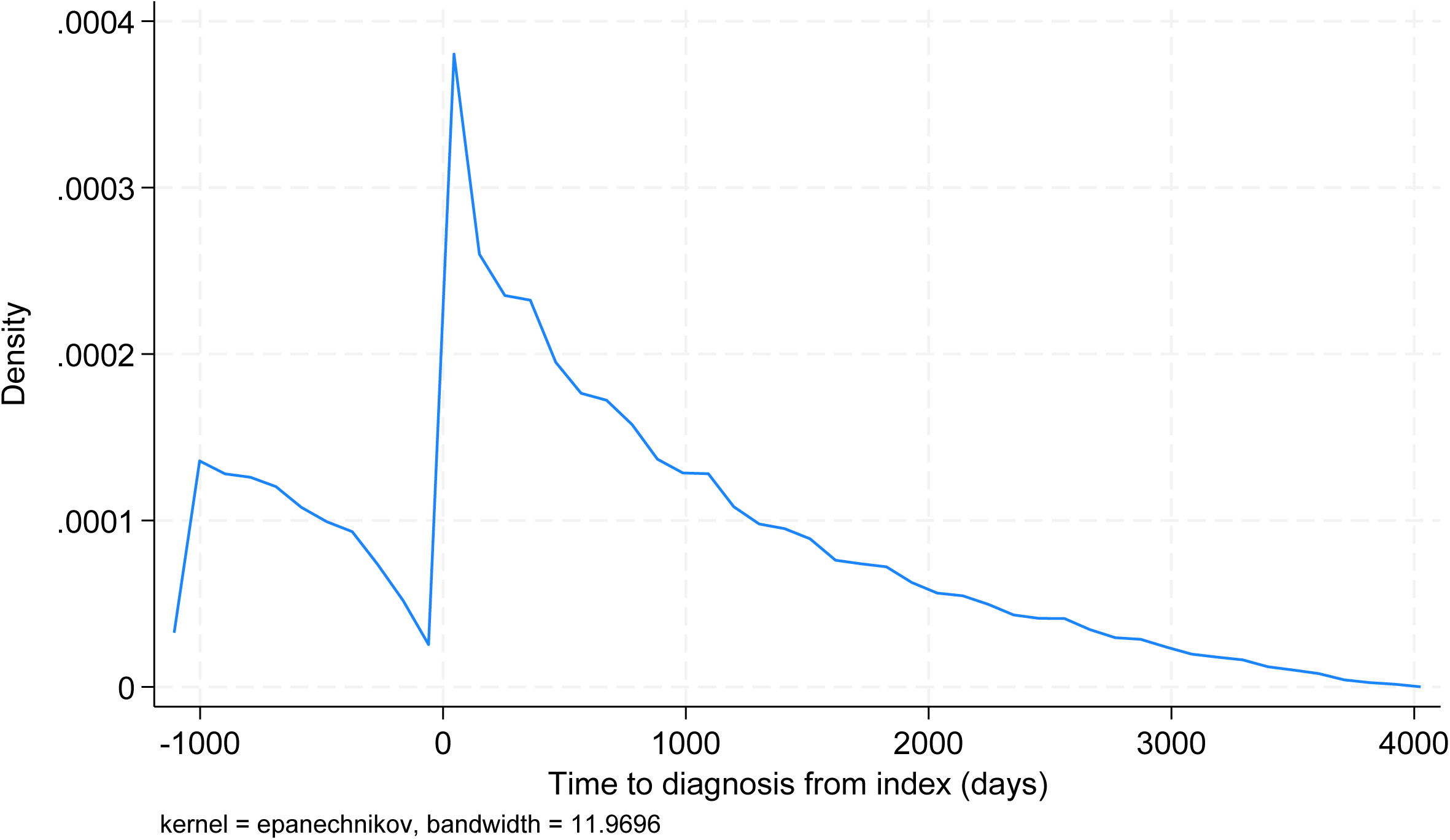
Kernel density plot of tonsillitis/sore throat episodes over the cohort period in reference to the index date.

**Supplementary table S1:**
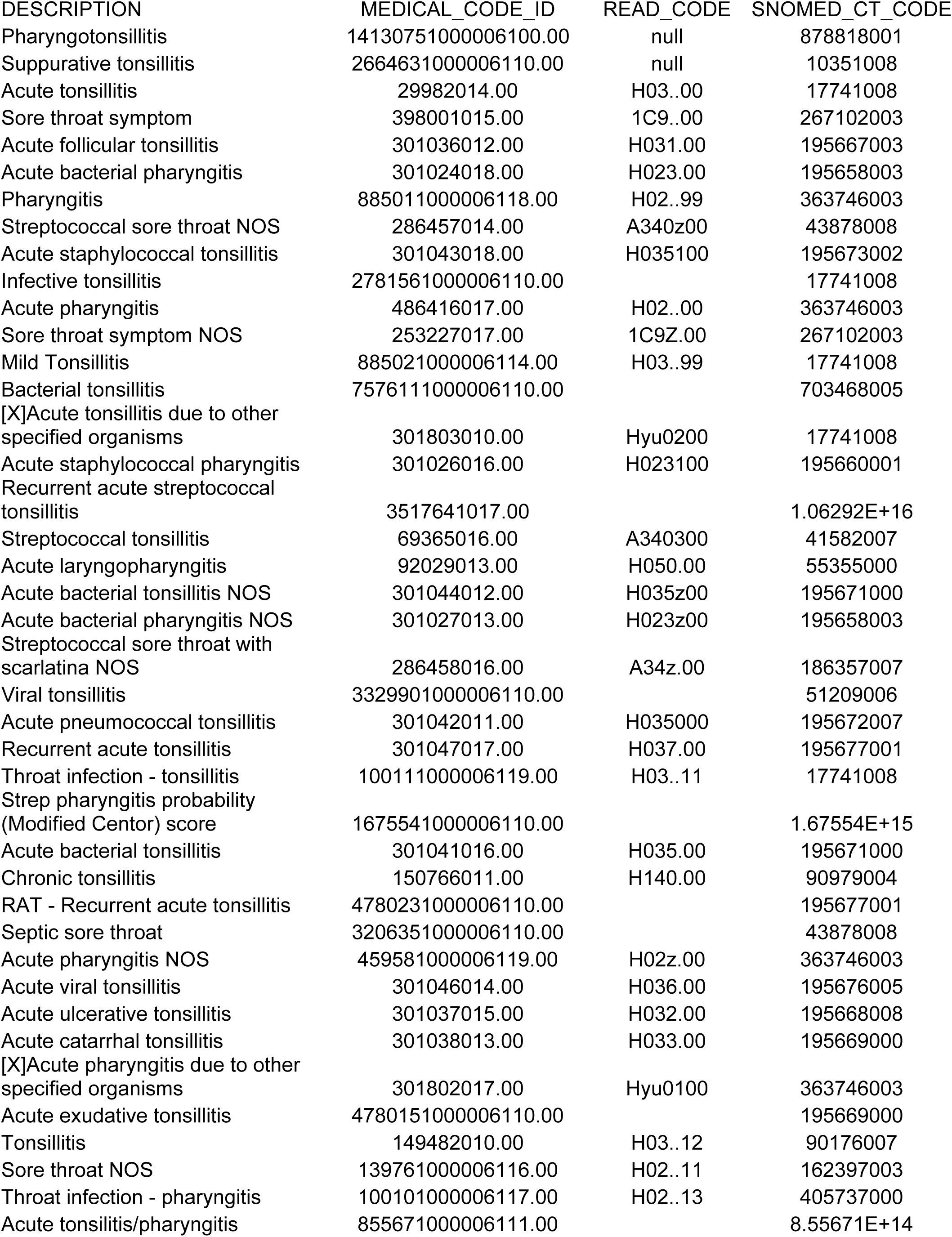
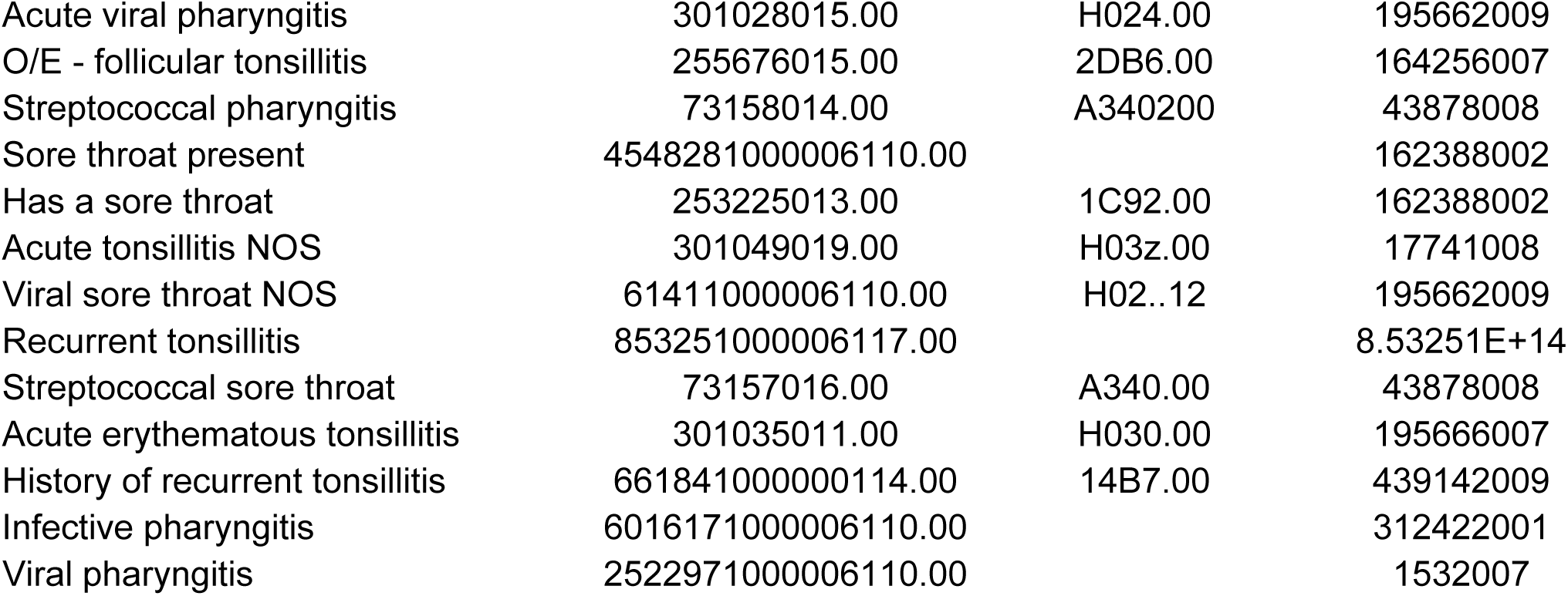
sore throat and tonsillitis codes.

| DESCRIPTION | MEDICAL_CODE_ID | READ_CODE | SNOMED_CT_CODE |
| --- | --- | --- | --- |
| Pharyngotonsillitis | 14130751000006100.00 | null | 878818001 |
| Suppurative tonsillitis | 2664631000006110.00 | null | 10351008 |
| Acute tonsillitis | 29982014.00 | H03..00 | 17741008 |
| Sore throat symptom | 398001015.00 | 1C9..00 | 267102003 |
| Acute follicular tonsillitis | 301036012.00 | H031.00 | 195667003 |
| Acute bacterial pharyngitis | 301024018.00 | H023.00 | 195658003 |
| Pharyngitis | 885011000006118.00 | H02..99 | 363746003 |
| Streptococcal sore throat NOS | 286457014.00 | A340z00 | 43878008 |
| Acute staphylococcal tonsillitis | 301043018.00 | H035100 | 195673002 |
| Infective tonsillitis | 2781561000006110.00 |  | 17741008 |
| Acute pharyngitis | 486416017.00 | H02..00 | 363746003 |
| Sore throat symptom NOS | 253227017.00 | 1C9Z.00 | 267102003 |
| Mild Tonsillitis | 885021000006114.00 | H03..99 | 17741008 |
| Bacterial tonsillitis | 7576111000006110.00 |  | 703468005 |
| [X]Acute tonsillitis due to other specified organisms | 301803010.00 | Hyu0200 | 17741008 |
| Acute staphylococcal pharyngitis | 301026016.00 | H023100 | 195660001 |
| Recurrent acute streptococcal tonsillitis | 3517641017.00 |  | 1.06292E+16 |
| Streptococcal tonsillitis | 69365016.00 | A340300 | 41582007 |
| Acute laryngopharyngitis | 92029013.00 | H050.00 | 55355000 |
| Acute bacterial tonsillitis NOS | 301044012.00 | H035z00 | 195671000 |
| Acute bacterial pharyngitis NOS | 301027013.00 | H023z00 | 195658003 |
| Streptococcal sore throat with scarlatina NOS | 286458016.00 | A34z.00 | 186357007 |
| Viral tonsillitis | 3329901000006110.00 |  | 51209006 |
| Acute pneumococcal tonsillitis | 301042011.00 | H035000 | 195672007 |
| Recurrent acute tonsillitis | 301047017.00 | H037.00 | 195677001 |
| Throat infection - tonsillitis | 100111000006119.00 | H03..11 | 17741008 |
| Strep pharyngitis probability (Modified Centor) score | 1675541000006110.00 |  | 1.67554E+15 |
| Acute bacterial tonsillitis | 301041016.00 | H035.00 | 195671000 |
| Chronic tonsillitis | 150766011.00 | H140.00 | 90979004 |
| RAT - Recurrent acute tonsillitis | 4780231000006110.00 |  | 195677001 |
| Septic sore throat | 3206351000006110.00 |  | 43878008 |
| Acute pharyngitis NOS | 459581000006119.00 | H02z.00 | 363746003 |
| Acute viral tonsillitis | 301046014.00 | H036.00 | 195676005 |
| Acute ulcerative tonsillitis | 301037015.00 | H032.00 | 195668008 |
| Acute catarrhal tonsillitis | 301038013.00 | H033.00 | 195669000 |
| [X]Acute pharyngitis due to other specified organisms | 301802017.00 | Hyu0100 | 363746003 |
| Acute exudative tonsillitis | 4780151000006110.00 |  | 195669000 |
| Tonsillitis | 149482010.00 | H03..12 | 90176007 |
| Sore throat NOS | 139761000006116.00 | H02..11 | 162397003 |
| Throat infection - pharyngitis | 100101000006117.00 | H02..13 | 405737000 |
| Acute tonsillitis/pharyngitis | 855671000006111.00 |  | 8.55671E+14 |
| Acute viral pharyngitis | 301028015.00 | H024.00 | 195662009 |
| O/E - follicular tonsillitis | 255676015.00 | 2DB6.00 | 164256007 |
| Streptococcal pharyngitis | 73158014.00 | A340200 | 43878008 |
| Sore throat present | 4548281000006110.00 |  | 162388002 |
| Has a sore throat | 253225013.00 | 1C92.00 | 162388002 |
| Acute tonsillitis NOS | 301049019.00 | H03z.00 | 17741008 |
| Viral sore throat NOS | 61411000006110.00 | H02..12 | 195662009 |
| Recurrent tonsillitis | 853251000006117.00 |  | 8.53251E+14 |
| Streptococcal sore throat | 73157016.00 | A340.00 | 43878008 |
| Acute erythematous tonsillitis | 301035011.00 | H030.00 | 195666007 |
| History of recurrent tonsillitis | 661841000000114.00 | 14B7.00 | 439142009 |
| Infective pharyngitis | 6016171000006110.00 |  | 312422001 |
| Viral pharyngitis | 2522971000006110.00 |  | 1532007 |

**Supplementary table S2:** Number people experiencing each number of episodes.

| Number of episodes | Tonsillitis | Sore throat | All episodes |
| --- | --- | --- | --- |
| 1 | 477,763 | 916,392 | 1,047,938 |
| 2 | 127,270 | 242,231 | 348,903 |
| 3 | 48,055 | 81,793 | 145,788 |
| 4 | 20,842 | 32,960 | 70,842 |
| 5 | 10,020 | 14,407 | 37,165 |
| 6 | 4,843 | 7,415 | 20,973 |
| 7 | 2,551 | 3,882 | 12,230 |
| 8 | 1,373 | 2,302 | 7,432 |
| 9 | 774 | 1,267 | 4,618 |
| 10+ | 1,104 | 2,712 | 8,815 |

**Supplementary table S3:** The proportion of people experiencing different number of episodes per year in the 3 years prior to and the three years after the baseline year along with mean episode rate per year according to the number of episodes in the baseline year.

| Episodes<br>in baseline<br>year | Year -1 |  |  |  |  |  |  |  | Year -2 |  |  |  |  |  |  |  | Year -3 |  |  |  |  |  |  |  | Mean episodes |  |  |
| --- | --- | --- | --- | --- | --- | --- | --- | --- | --- | --- | --- | --- | --- | --- | --- | --- | --- | --- | --- | --- | --- | --- | --- | --- | --- | --- | --- |
|  | 0 | 1 | 2 | 3 | 4 | 5 | 6 | >6 | 0 | 1 | 2 | 3 | 4 | 5 | 6 | >6 | 0 | 1 | 2 | 3 | 4 | 5 | 6 | >6 | Year<br>-1 | Year<br>-2 | Year<br>-3 |
| 1 | 99.1 | 0.7 | 0.1 | 0.0 | 0.0 | 0.0 | 0.0 | 0.0 | 94.52 | 5 | 0 | 0.01 | 0 | 0 | 0 | 0 | 91.3 | 8.7 | 0 | 0 | 0 | 0 | 0 | 0 | 0.01 | 0.06 | 0.09 |
| 2 | 83.5 | 15.62 | 0.7 | 0.14 | 0.03 | 0.01 | 0 | 0 | 84.37 | 10 | 5 | 0.21 | 0.04 | 0.01 | 0 | 0 | 83.95 | 6.9 | 9.15 | 0 | 0 | 0 | 0 | 0 | 0.18 | 0.22 | 0.25 |
| 3 | 75.53 | 13.21 | 10.46 | 0.57 | 0.17 | 0.05 | 0 | 0 | 74 | 10 | 10.34 | 5.37 | 0.34 | 0.09 | 0.01 | 0 | 76.49 | 6.29 | 7.76 | 9.47 | 0 | 0 | 0 | 0 | 0.37 | 0.48 | 0.50 |
| 4 | 69.11 | 12.81 | 9.02 | 8.17 | 0.57 | 0.23 | 0.05 | 0.05 | 65 | 9.57 | 10.41 | 10 | 4.22 | 0.37 | 0.07 | 0 | 70.78 | 4.32 | 5.8 | 9.22 | 9.89 | 0 | 0 | 0 | 0.59 | 0.80 | 0.83 |
| 5 | 62.67 | 13.54 | 8.41 | 7.9 | 6.36 | 0.51 | 0.31 | 0.31 | 59.49 | 7.38 | 10.05 | 8.82 | 10.05 | 3.08 | 1.03 | 0 | 65.33 | 3.59 | 4.31 | 7.38 | 9.85 | 9.54 | 0 | 0 | 0.86 | 1.16 | 1.21 |
| 6 | 52.49 | 15.71 | 7.28 | 6.13 | 6.9 | 6.13 | 1.53 | 3.83 | 55.56 | 4.98 | 6.51 | 6.51 | 7.28 | 10.73 | 6.51 | 1.92 | 64.37 | 4.98 | 2.68 | 3.07 | 4.6 | 8.81 | 11.49 | 0 | 1.44 | 1.75 | 1.51 |
| >6 | 41.43 | 5.71 | 5 | 5 | 7.14 | 5 | 15 | 15.71 | 50 | 1.43 | 3.57 | 5 | 5 | 5 | 7.86 | 22.14 | 54.3 | 3.6 | 0.7 | 0.7 | 2.1 | 2.1 | 11.4 | 25.0 | 2.94 | 2.79 | 2.84 |

| Episodes<br>in baseline<br>year | Year 1 |  |  |  |  |  |  |  | Year 2 |  |  |  |  |  |  |  | Year 3 |  |  |  |  |  |  |  | Mean episodes |  |  |
| --- | --- | --- | --- | --- | --- | --- | --- | --- | --- | --- | --- | --- | --- | --- | --- | --- | --- | --- | --- | --- | --- | --- | --- | --- | --- | --- | --- |
|  | 0 | 1 | 2 | 3 | 4 | 5 | 6 | >6 | 0 | 1 | 2 | 3 | 4 | 5 | 6 | >6 | 0 | 1 | 2 | 3 | 4 | 5 | 6 | >6 | Year<br>1 | Year<br>2 | Year<br>3 |
| 1 | 87.1 | 11.4 | 1.3 | 0.2 | 0.0 | 0.0 | 0.0 | 0.0 | 84 | 14 | 2 | 0.25 | 0.04 | 0.01 | 0 | 0 | 91.85 | 7.1 | 0.87 | 0.14 | 0.02 | 0.01 | 0 | 0 | 0.15 | 0.18 | 0.09 |
| 2 | 74.84 | 19.99 | 4.13 | 0.83 | 0.17 | 0.03 | 0.01 | 0.01 | 70.2 | 24 | 4 | 0.86 | 0.18 | 0.04 | 0.01 | 0 | 83.4 | 13.4 | 2.57 | 0.48 | 0.1 | 0.02 | 0.01 | 0 | 0.32 | 0.37 | 0.21 |
| 3 | 62.44 | 26.44 | 8.2 | 2.12 | 0.6 | 0.13 | 0.05 | 0.02 | 58 | 31 | 8.16 | 2.06 | 0.53 | 0.16 | 0.03 | 0 | 75.46 | 18.3 | 4.6 | 1.24 | 0.33 | 0.06 | 0.03 | 0 | 0.53 | 0.57 | 0.33 |
| 4 | 50.94 | 28.63 | 12.79 | 5.18 | 1.46 | 0.62 | 0.3 | 0.09 | 49 | 33.7 | 11.19 | 3.36 | 1.44 | 0.53 | 0.27 | 0 | 69.43 | 21.2 | 6.37 | 2.08 | 0.66 | 0.21 | 0.02 | 0.1 | 0.81 | 0.77 | 0.44 |
| 5 | 41.74 | 30.05 | 14.26 | 6.97 | 3.79 | 1.54 | 1.44 | 0.21 | 41.85 | 33.1 | 13.95 | 4.31 | 3.69 | 1.44 | 1.03 | 1 | 62.87 | 19.6 | 10.4 | 3.49 | 2.26 | 0.92 | 0.21 | 0.3 | 1.13 | 1.07 | 0.68 |
| 6 | 31.42 | 21.46 | 15.71 | 11.9 | 3.07 | 7.66 | 4.6 | 4.21 | 42.53 | 23.4 | 11.88 | 6.51 | 4.21 | 4.98 | 2.3 | 4.21 | 62.07 | 14.9 | 8.43 | 5.75 | 2.68 | 2.68 | 0.77 | 2.7 | 2.00 | 1.55 | 0.97 |
| >6 | 17.14 | 16.43 | 7.14 | 7.86 | 6.43 | 7.86 | 12.1 | 25 | 31.43 | 14.3 | 7.86 | 7.86 | 2.14 | 5.71 | 7.14 | 23.57 | 44.3 | 12.9 | 6.4 | 5.7 | 5.0 | 7.9 | 5.0 | 12.9 | 3.81 | 3.19 | 2.26 |

**Supplementary table S4:** Antibiotics used to treat episodes of tonsillitis or sore throat.

| Episode type | Antibiotic used (N(%)) |  |  |  |  |  |
| --- | --- | --- | --- | --- | --- | --- |
|  | None | Amoxicillin | Clarithromycin | Clindamycin | Erythromycin | Penicillin V |
| Tonsillitis | 196,620 (18.1) | 51,501 (4.7) | 65,827 (60.1) | 117 (0.01) | 47,658 (4.4) | 726,029 (66.7) |
| Sore throat | 1,199,515 (60.3) | 200,230 (10.1) | 71,757 (3.6) | 84 (0.00) | 57,908 (2.9) | 458,882 (23.1) |

**Supplementary table S5:** Number of patients referred to ENT and the time to referral from index, along with the number of patients who underwent tonsillectomy and the time to tonsillectomy from index for each subgroup population.

| Group | N | ENT Ref N(%<br>of group) | Days to ENT referral<br>from index (mean(SD)) | Tonsillectomy<br>N(% of<br>group) | Days to tonsillectomy from index<br>(mean(SD)) |
| --- | --- | --- | --- | --- | --- |
| Full cohort | 4,453,538 | 368,187 (8.3) | 1080 (921) | 21,869 (0.49) | 737 (731) |
| Exposed | 1,704,704 | 215,097<br>(12.6) | 985 (919) | 20,507 (1.2) | 721 (723) |
| Unexposed | 2,748,834 | 153,090 (5.6) | 1215 (906) | 1,362 (0.05) | 972 (811) |
| Paradise<br>positive at index | 242 | 78 (32.2) | 1,296 (1,008) | 15 (6.2) | 581 (835) |
| Paradise<br>positive at any<br>time | 40,427 | 14,481 (35.8) | 814 (873) | 5,629 (13.9) | 775 (703) |

**Supplementary table S6:** Multinomial logistic regression model for tonsillectomy with clustering at practice ID.

|  |  | Odds ratio | p | 95% conf. interval |  |
| --- | --- | --- | --- | --- | --- |
| Paradise positive |  | 27.523 | <0.005 | 26.332 | 28.769 |
| Age category (ref 16-25) | 26-35 | 0.459 | <0.005 | 0.440 | 0.480 |
|  | 36-45 | 0.268 | <0.005 | 0.255 | 0.283 |
|  | 46-55 | 0.142 | <0.005 | 0.133 | 0.153 |
|  | 56-65 | 0.093 | <0.005 | 0.085 | 0.103 |
|  | 66-75 | 0.061 | <0.005 | 0.054 | 0.070 |
|  | 76+ | 0.025 | <0.005 | 0.019 | 0.033 |
| Sex (ref male) | Female | 1.183 | <0.005 | 1.136 | 1.232 |
| Ethnicity (ref White) | Black | 0.678 | <0.005 | 0.602 | 0.765 |
|  | Asian | 0.677 | <0.005 | 0.626 | 0.733 |
|  | Mixed | 0.890 | 0.328 | 0.706 | 1.123 |
|  | Other | 0.893 | 0.147 | 0.767 | 1.041 |
|  | Missing | 0.850 | <0.005 | 0.805 | 0.897 |
| IMD Quintile (ref 1st, least deprived) | 2nd | 0.952 | 0.114 | 0.895 | 1.012 |
|  | 3rd | 0.896 | 0.001 | 0.841 | 0.955 |
|  | 4th | 0.855 | <0.005 | 0.801 | 0.913 |
|  | 5th | 0.798 | <0.005 | 0.746 | 0.855 |
|  | Missing | 0.839 | <0.005 | 0.777 | 0.906 |
| Region (ref East Midlands) | East of England | 1.074 | 0.480 | 0.881 | 1.310 |
|  | London | 0.983 | 0.843 | 0.833 | 1.161 |
|  | North East | 1.182 | 0.086 | 0.977 | 1.431 |
|  | North West | 1.185 | 0.041 | 1.007 | 1.394 |
|  | Northern Ireland | 1.824 | <0.005 | 1.328 | 2.507 |
|  | South East | 1.021 | 0.807 | 0.867 | 1.202 |
|  | South West | 0.935 | 0.447 | 0.785 | 1.113 |
|  | West Midlands | 0.965 | 0.671 | 0.818 | 1.138 |
|  | Yorkshire and The Humber | 1.004 | 0.972 | 0.820 | 1.228 |
| Smoking status (ref never) | Ex-smoker | 1.120 | <0.005 | 1.072 | 1.171 |
|  | Current smoker | 1.000 | 0.992 | 0.956 | 1.045 |
|  | Missing | 0.699 | <0.005 | 0.593 | 0.823 |
| BMI (per kg/m<sup>2</sup>) |  | 1.021 | <0.005 | 1.019 | 1.024 |
| Constant |  | 0.006 | <0.005 | 0.005 | 0.007 |

**Supplementary table S7:** Multinomial logistic regression model for ENT referral (any and within 180d of episode) with clustering at practice ID.

|  |  | Any ENT referral |  |  |  | ENT Referrals within 180d of episode |  |  |  |
| --- | --- | --- | --- | --- | --- | --- | --- | --- | --- |
|  |  | Odds ratio | P value | [95% conf. interval] |  | Odds ratio | P value | [95% conf. interval] |  |
| Paradise positive |  | 4.62 | <0.005 | 3.36 | 6.34 | 8.95 | <0.005 | 6.01 | 13.32 |
| Age cate (ref 16-25 years) | 26-35 | 0.96 | <0.005 | 0.95 | 0.98 | 0.65 | <0.005 | 0.63 | 0.66 |
|  | 36-45 | 1.25 | <0.005 | 1.23 | 1.27 | 0.63 | <0.005 | 0.62 | 0.65 |
|  | 46-55 | 1.52 | <0.005 | 1.50 | 1.55 | 0.73 | <0.005 | 0.71 | 0.76 |
|  | 56-65 | 1.78 | <0.005 | 1.75 | 1.81 | 0.81 | <0.005 | 0.78 | 0.83 |
|  | 66-75 | 1.93 | <0.005 | 1.90 | 1.97 | 0.89 | <0.005 | 0.86 | 0.92 |
|  | 76+ | 1.76 | <0.005 | 1.72 | 1.79 | 0.86 | <0.005 | 0.83 | 0.89 |
| Sex (ref male) | Female | 1.08 | <0.005 | 1.07 | 1.09 | 0.98 | <0.005 | 0.96 | 0.99 |
| Ethnicity (ref white) | Black | 0.81 | <0.005 | 0.79 | 0.83 | 0.87 | <0.005 | 0.83 | 0.91 |
|  | Asian | 1.12 | <0.005 | 1.11 | 1.14 | 1.20 | <0.005 | 1.17 | 1.24 |
|  | Mixed | 0.88 | <0.005 | 0.83 | 0.93 | 1.11 | 0.048 | 1.00 | 1.23 |
|  | Other | 0.93 | <0.005 | 0.90 | 0.97 | 0.98 | 0.596 | 0.91 | 1.06 |
|  | Missing | 0.66 | <0.005 | 0.65 | 0.67 | 0.72 | <0.005 | 0.70 | 0.75 |
| IMD (ref 1st) | 2nd | 0.97 | <0.005 | 0.96 | 0.99 | 1.00 | 0.791 | 0.97 | 1.03 |
|  | 3rd | 0.97 | <0.005 | 0.95 | 0.98 | 0.98 | 0.153 | 0.95 | 1.01 |
|  | 4th | 0.97 | <0.005 | 0.95 | 0.98 | 0.96 | 0.012 | 0.93 | 0.99 |
|  | 5th | 0.97 | 0.003 | 0.96 | 0.99 | 0.98 | 0.311 | 0.95 | 1.02 |
|  | missing | 0.99 | 0.524 | 0.96 | 1.02 | 0.99 | 0.574 | 0.94 | 1.04 |
| Area (ref East Midlands) | East of England | 1.34 | <0.005 | 1.17 | 1.53 | 1.26 | <0.005 | 1.07 | 1.47 |
|  | London | 1.21 | <0.005 | 1.09 | 1.34 | 1.15 | 0.032 | 1.01 | 1.30 |
|  | North East | 1.43 | <0.005 | 1.27 | 1.62 | 1.36 | <0.005 | 1.17 | 1.57 |
|  | North West | 1.51 | <0.005 | 1.36 | 1.67 | 1.47 | <0.005 | 1.29 | 1.66 |
|  | Northern Ireland | 2.11 | <0.005 | 1.71 | 2.59 | 1.71 | <0.005 | 1.33 | 2.20 |
|  | South East | 1.15 | 0.009 | 1.04 | 1.28 | 1.06 | 0.346 | 0.94 | 1.21 |
|  | South West | 0.99 | 0.929 | 0.89 | 1.11 | 0.91 | 0.182 | 0.80 | 1.04 |
|  | West Midlands | 1.11 | 0.053 | 1.00 | 1.23 | 1.06 | 0.391 | 0.93 | 1.20 |
|  | Yorkshire and The Humber | 1.30 | <0.005 | 1.14 | 1.49 | 1.22 | 0.013 | 1.04 | 1.43 |
| Smoking status | Ex-smoker | 1.12 | <0.005 | 1.11 | 1.13 | 1.15 | <0.005 | 1.13 | 1.17 |
|  | Current smoker | 0.95 | <0.005 | 0.94 | 0.96 | 0.97 | 0.006 | 0.95 | 0.99 |
|  | Missing | 0.58 | <0.005 | 0.54 | 0.61 | 0.57 | <0.005 | 0.51 | 0.64 |
| BMI (per kg/m<sup>2</sup>) |  | 1.00 | <0.005 | 1.00 | 1.00 | 1.01 | <0.005 | 1.01 | 1.01 |
| Constant |  | 0.05 | <0.005 | 0.05 | 0.06 | 0.02 | <0.005 | 0.02 | 0.02 |

**Supplementary table S8:** Time-to-event analysis for episode of tonsillitis/sore throat from the index episode considering multiple events using the Anderson-Gill Cox model.

| Variable |  | Hazard ratio | 95% CI |  | P value |
| --- | --- | --- | --- | --- | --- |
| Number of episodes in the 3 years prior to index | 1 | 1.83 | <0.005 | 1.77 | 1.89 |
|  | 2 | 2.49 | <0.005 | 2.38 | 2.61 |
|  | 3 | 3.42 | <0.005 | 3.18 | 3.68 |
|  | 4 | 4.03 | <0.005 | 3.60 | 4.51 |
|  | 5 | 5.39 | <0.005 | 4.55 | 6.40 |
|  | 6 | 5.11 | <0.005 | 4.24 | 6.16 |
|  | 7 | 7.78 | <0.005 | 5.68 | 10.66 |
|  | 8 | 7.68 | <0.005 | 4.56 | 12.92 |
|  | 9 | 8.87 | <0.005 | 4.92 | 16.02 |
|  | 10 | 6.12 | <0.005 | 5.85 | 6.40 |
|  | 11+ | 24.64 | <0.005 | 16.27 | 37.30 |
| Age category (ref 16-25) | 26-35 | 0.78 | <0.005 | 0.75 | 0.80 |
|  | 36-45 | 0.58 | <0.005 | 0.57 | 0.60 |
|  | 46-55 | 0.50 | <0.005 | 0.48 | 0.52 |
|  | 56-65 | 0.45 | <0.005 | 0.44 | 0.48 |
|  | 66-75 | 0.45 | <0.005 | 0.42 | 0.47 |
|  | 76+ | 0.42 | <0.005 | 0.39 | 0.45 |
| Sex (ref male) |  | 1.39 | <0.005 | 1.36 | 1.42 |
| Ethnicity (ref White) | Black | 0.95 | 0.062 | 0.89 | 1.00 |
|  | Asian | 1.24 | <0.005 | 1.20 | 1.29 |
|  | Mixed | 1.10 | 0.195 | 0.95 | 1.26 |
|  | Other | 0.90 | 0.02 | 0.82 | 0.98 |
|  | Missing | 0.96 | 0.016 | 0.93 | 0.99 |
| IMD quintile (ref 1st) | 2 | 1.00 | 0.808 | 0.96 | 1.03 |
|  | 3 | 1.04 | 0.03 | 1.00 | 1.08 |
|  | 4 | 1.05 | 0.01 | 1.01 | 1.09 |
|  | 5 | 1.08 | <0.005 | 1.04 | 1.12 |
|  | Missing | 1.05 | 0.016 | 1.01 | 1.08 |
| Smoking status (ref none/never) | Ex-smoker | 1.00 | 0.71 | 0.98 | 1.03 |
|  | Current smoker | 1.01 | 0.644 | 0.98 | 1.03 |
|  | Missing | 1.04 | 0.517 | 0.92 | 1.17 |
| BMI (per kg/m<sup>2</sup>) |  | 1.01 | <0.005 | 1.01 | 1.01 |

**Supplementary table S9:** Breakdown of episodes including antibiotic defined episodes along with the co-prescribed antibiotics.

| Episode type | Group (N(%)) |  | Antibiotic used (N(%)) |  |  |  |  |  | Total |
| --- | --- | --- | --- | --- | --- | --- | --- | --- | --- |
|  | Unexposed | Exposed | None | Amoxicillin | Clarithromycin | Clindamycin | Erythromycin | Penicillin V |  |
| Tonsillitis | 0 | 1,083,678<br>(32.3) | 192,256<br>(17.8) | 51,470<br>(4.6) | 65,927<br>(6.1) | 117<br>(0.01) | 47,688<br>(4.4) | 726,217<br>(67.0) | 1,083,675 |
| Sore throat | 0 | 1,984,282<br>(59.2) | 1,195,123<br>(60.2) | 200,298<br>(10.1) | 71,781<br>(3.6) | 84<br>(0.0) | 57,950<br>(2.9) | 459,046<br>(23.1) | 1,984,282 |
| Antibiotic episode | 204,688<br>(100) | 284,634<br>(8.5) | 0 | 0 | 0 | 0 | 0 | 489,322<br>(100) | 489,322 |
| Total | 204,688 | 3,352,591 | 1,387,379<br>(39.0) | 251,768<br>(7.1) | 137,708<br>(3.9) | 201<br>(0.01) | 105,638<br>(3.0) | 1,674,585<br>(47.1) | 3,557,279 |

*Supplementary material S1: Exploratory analysis examining antibiotic defined episodes to estimate the proportion of episodes that are uncoded.*

1,469,897 episodes treated with penicillin V in the exposed representing 44% of all coded episodes.

Assuming the unexposed population have the same rate of tonsillitis and sore throat episodes, and the same proportion of episodes treated by penicillin V as the exposed arm, there would be 465,200 (204,688/0.44) episodes in the unexposed arm that haven’t been identified. This would be 465,200 out of 3,817,791 episodes in total. This equates to 6.8% of episodes not identified through codes. The episodes used in the main analysis (only coded episodes) therefore make up, assuming all the antibiotic defined episodes are accurate, 76% of all episodes.

